# Dendrite: A Structured, Accessible, and Queryable Pathology Search Database for Streamlined Experiment Planning

**DOI:** 10.1101/2023.09.09.23295302

**Authors:** Yunrui Lu, Robert Hamilton, Jack Greenberg, Gokul Srinivasan, Parth Shah, Sarah Preum, Jason Pettus, Louis Vaickus, Joshua Levy

## Abstract

Pathology reports contain vital information, yet a significant portion of this data remains underutilized in electronic medical record systems due to the unstructured and varied nature of reporting. Although synoptic reporting has introduced reporting standards, the majority of pathology text remains free-form, necessitating additional processing to enable accessibility for research and clinical applications. This paper presents Dendrite, a web application designed to enhance pathology research by providing intelligent search capabilities and streamlining the creation of study cohorts. Leveraging expert knowledge and natural language processing algorithms, Dendrite converts free-form pathology reports into structured formats, facilitating easier querying and analysis. Using a custom Python script, Dendrite organizes pathology report data, enabling record linkages, text searches, and structured drop-down menus for information filtering and integration. A companion web application enables data exploration and export, showcasing its potential for further analysis and research. Dendrite, derived from existing laboratory information systems, outperforms existing implementations in terms of speed, responsiveness, and flexibility. With its efficient search functionality and support for clinical research and quality improvement efforts in the pathology field, Dendrite proves to be a valuable tool for pathologists. Future enhancements encompass user management integration, integration of natural language processing and machine learning to enhance structured reporting capabilities and seamless integration of Dendrite with the vast repository of genomics and imaging data.

## Introduction

Pathology, derived from the word *pathos*, is the study of disease. Disease progression is largely characterized by the use of increasingly sophisticated histological and molecular assays. Pathological examination of these assays plays a critical role in diagnosis, prognostication, and treatment, potentially enhancing prevention and screening efforts ^1^. For instance, histological and molecular information often serves as the baseline and endpoint of drug trials for clinical oncology applications ^2^. Pathology reports play a crucial role in capturing the clinical narrative reflecting these assessments, encompassing vital information related to diagnosis, prognosis, and specimen processing. Traditional approaches in natural language processing (NLP) have employed rule-based or machine-learning analytics to extract valuable insights from textual patterns contained in these reports, enabling clinical endpoints and biomarker information to be derived from these reports ^3^.

Leveraging the wealth of information in pathology reports for large-scale databases is costly and labor-intensive, but if harnessed, has the potential to contribute significantly to comprehensive cancer registries, enabling more precise population-level studies and identification of novel associations between clinical features and outcomes. The subjective nature and use of nonstandard mapping terminology in anatomic pathology reports limit their interoperability with electronic medical record (EMR) systems– many efforts have been taken to further structure this information into digestible queryable formats such as the migration of synoptic reporting information into EPIC Beaker ^4,5^.

Structuring this data into more standardized formats has the potential to improve the utilization of pathological reporting information. Such resources could be readily pooled into larger electronic health record systems such as EPIC, which could spur future research/clinical applications. Currently, many clinical research studies still rely on manual chart review, which is often inefficient and prone to error. To overcome these challenges, there is a growing need for advanced database management tools that can efficiently store, manage, and analyze pathology data for research purposes. The use of database resources can enhance the efficiency of clinical research and quality improvement planning. Collaborating with a domain expert pathologist in developing these databases can ensure that the captured information is pertinent and applicable to the work of various diagnostic subspecialties. Furthermore, several recent studies have sought to leverage restructured pathology text reports to perform various classification (e.g., study case complexity for reimbursement and RVUs) and information extraction tasks (e.g., extracting gross/histo-morphology). Many of these algorithms operate on unstructured free text ^6,7^.

Our pathology department at a mid-sized academic center, is transitioning from an Oracle Cerner laboratory information system (LIS) to EPIC Beaker LIS. We were motivated by previous attempts to create business intelligence systems that could swiftly retrieve structured and unstructured pathology reporting data ^8^. As a result, we designed our own in-house solution with enhanced search capabilities, called Dendrite. Dendrite leverages an internal pathology reporting database structured from pathology notes.

Dendrite refers to both the pathology reporting database, the codebase used to generate this database as well as its front facing web interface. This database encompasses nearly 1 million pathology reports, extracted from 2008 to 2022 across various diagnostic subspecialties. Dendrite, designed by pathologists for pathologists, employs traditional extraction methods based on expert domain knowledge to capture reporting information such as staining results and procedural codes along with nearly one hundred additional reporting fields. Dendrite allows for the arbitrary combination of multiple search criteria spanning nearly all facets of the pathology search, with capabilities similar to systems like Cerner, allowing for the rapid display of disparate information sources. The platform was developed to enable a real-time human-computer web application interface that supports interactive search, filtering and aggregation. Reports can be viewed through an output display table that can be edited in real-time and exported. Dendrite has been integrated with the Dartmouth Cloud or Augmet, an AWS cloud resource that seamlessly integrates with the electronic health record (EHR) system ^9^. This integration has further improved Dendrite’s capabilities for imaging and genomics applications, enabling swift querying and exporting of genomics and imaging data in addition to a wide array of pathology reporting fields.

In the future, Dendrite can be used to supplement innovative cancer screening and surveillance approaches, enhance patient outcomes, facilitate the rapid prototyping and development of advanced machine learning algorithms for text, genomics, and imaging, and broaden our understanding of cancer epidemiology ^10–12^. The aim of this study is to demonstrate the functionality of this search tool. Subsequent studies will focus on documenting its implementation.

## Methods

### Data Collection and Development/Description of Dendrite Database

After IRB approval, we developed a set of custom Python scripts to process ten years worth of pathology reporting data, corresponding to 749,136 reports from the end of 2011 to 2021. We adopted an extract, transform, load (ETL) framework that used regex for report deidentification and stain dictionaries for the accurate reporting of 738 stains. It also parsed and delineated different report sections (e.g., dividing diagnostic text by specimen source and free text).

Reporting information was reorganized into the following tables:

- Stain orders and results
- Slide imaging information
- Cytology preparation methods
- Cytological findings
- Parsed and free diagnostic text
- Parsed and free discussion text
- Flow cytometry results
- Synoptics reporting
- Clinical history
- HPV results (cytology)
- Specimen adequacy
- Gross specimen reports
- Molecular findings
- Additional relevant anatomic pathology information (e.g., practicing pathologist, outside consult, EDTA, subspecialty identifiers, addendum, etc.)

**Supplementary Table 1** provides the schema / reporting fields for each table. This information can be filtered and joined through record linkages, text searches and structured drop-down menus (e.g., subsetting by stain). The dendrite database can be readily updated through automatic export of Cerner stored pathology reports and passing these reports through the Python script. All reporting fields were deidentified using the Python script that uses regex to remove information using a database of tens of thousands of first and last names. Dates were stripped although a custom index date was stored which allows for the recovery of this information.

This pathology database serves as the backend for an accompanying web-based tool that can query this information. A web application was developed using Plotly Dash ^13^, a Python tool that generates interactive websites– this interactive web application allows for collaborating pathologists to explore this database through multiple search tables, to be described in subsequent sections. We hosted this database and accompanying web application using an Amazon Web Services (AWS) instance for internal access.

### Web Application Description

### Multiple Search Criteria

The Dendrite web application layout was inspired by the web design of NCBI’s search tool and is divided into two sections (**Figure 1**): a) a tool for constructing filters to query the database (left panel) and b) a tool to explore the queried results and browse/export rapidly compiled information tables (right panel). The left panel is vertically scrollable to allow the addition of multiple filters while the right panel is static. Several buttons have been added which provide detailed instructions (a dynamically extending instructions tab on the right-hand side for additional usage details) for operating the application and providing further description on the database via a popup (**Figure 1A**). Below the instruction buttons are various buttons which control the addition of report database filters (i.e., Add/Remove filter) and the subunits of the filter, which allow for the selection of reporting criteria from multiple search tables (**Figure 1A**). By clicking on the Add Filter, physicians can add multiple filter units in order to build multiple search conditions.

**Figure 1:**
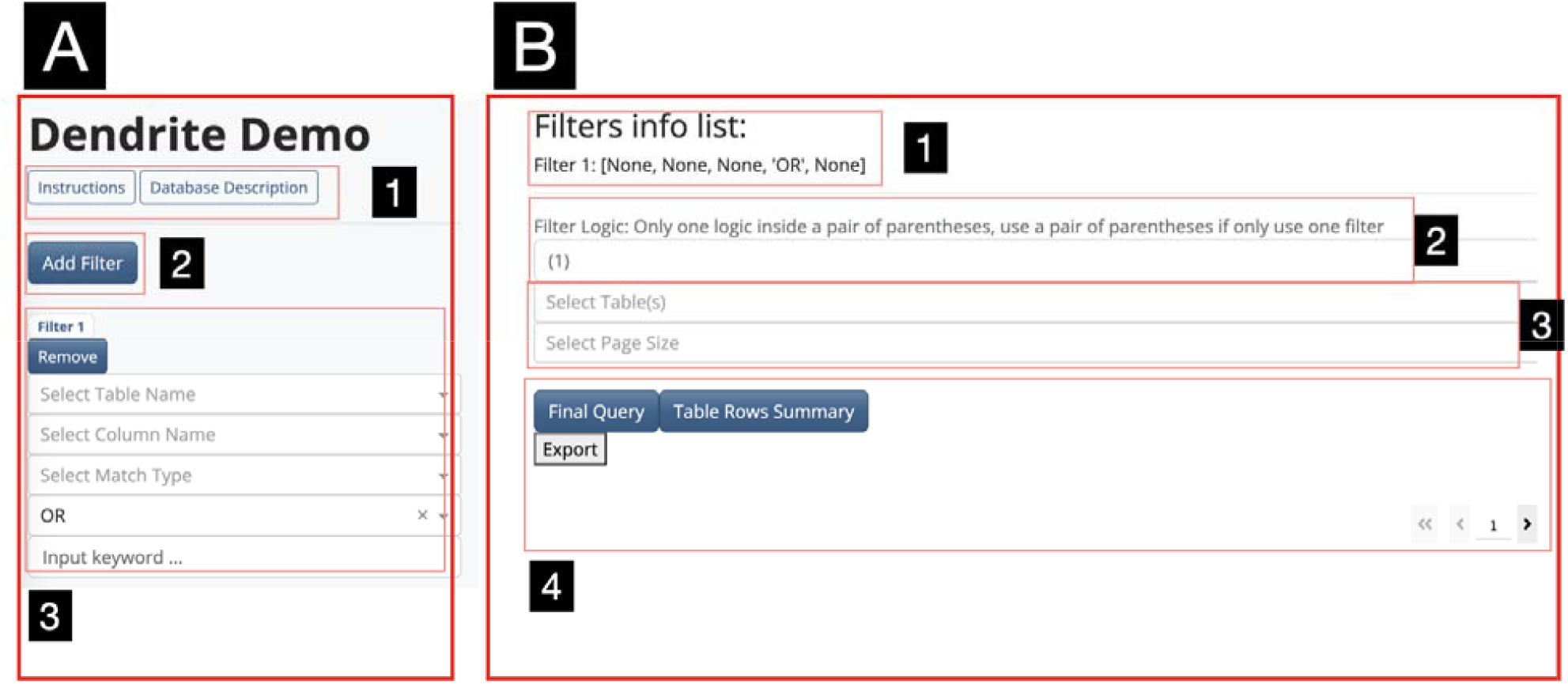
Dendrite Initial Interface: **A)** Filters component. (1) Instruction button and database description button. (2) Adding filters button. (3) Filter unit. **B)** Query component. (1) Filter list information display. (2) Logic statement input. (3) Target table(s) selection and page size selection. (4) Table query, information summary and display.

#### Description of Filter

Filters are specified by the user to query a *table* (e.g., all stains contained in stains_full_safe) for a specific field (e.g., stains as selected from stain_name) (**Figure 2**). The user is then prompted via an *input keyword* argument to enter free text to query the table field with (e.g., searching for carcinoma) should the field contain free text, else the user can select from a list of unique values from a dropdown menu that dynamically suggests these values based on an initial input. Physicians can either choose an individual term from a dropdown menu or input any keyword. Dendrite will then continue to use this keyword to generate further accurate suggestions. Physicians can, again, choose one of the suggestions or input any additional keywords to yield the final result. Furthermore, users can select whether query text from the selected field should exactly match or contain the keyword (the latter option produces more flexible searches at the expense of specificity). The creation of a filter will select all associated pathology reports by unique identifiers.

**Figure 2:**
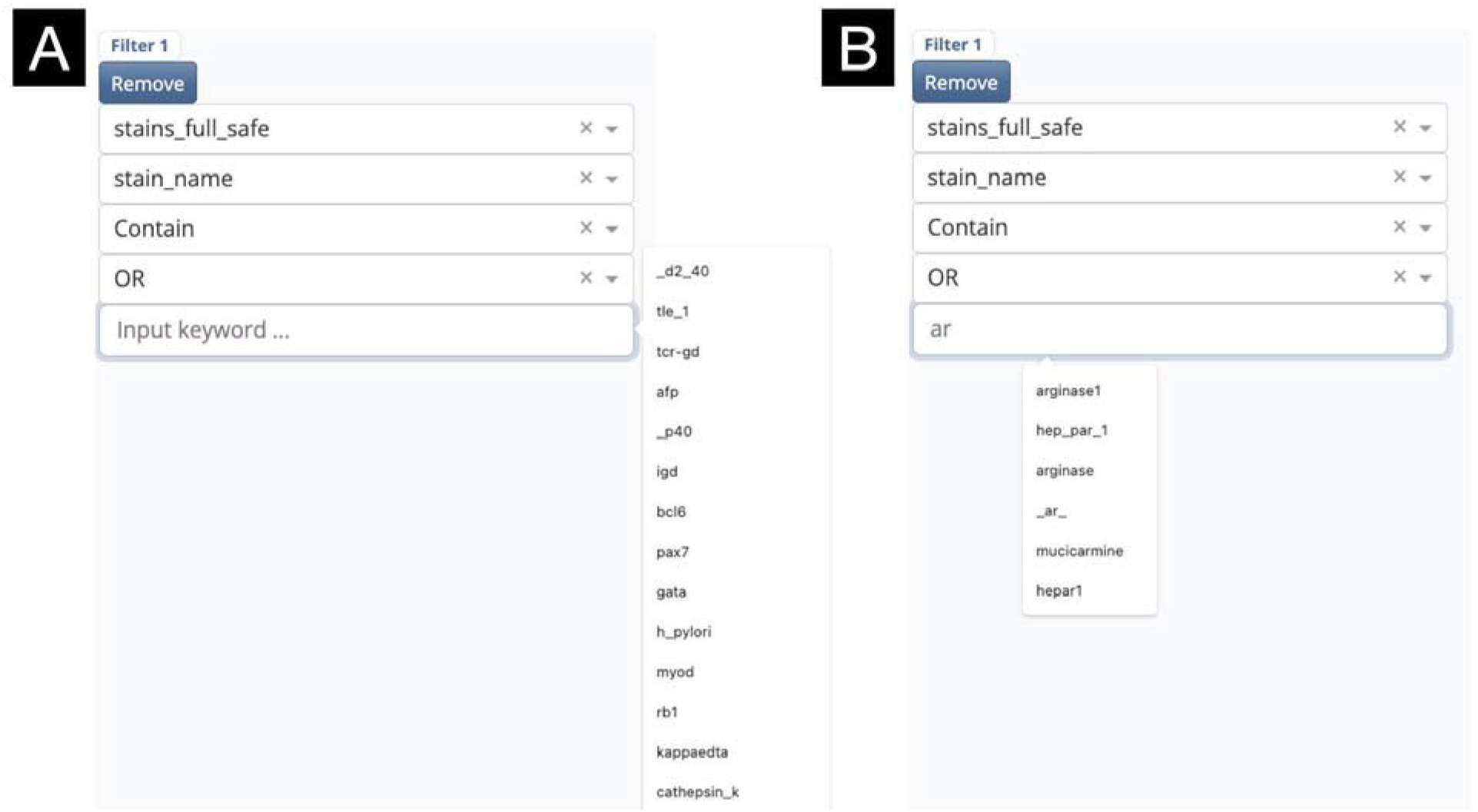
Keyword Input Prompts: **(A)** Initial prompts. **(B)** Prompts automatically refined through user input

### Adjustable Search Logic

Filters are indexed (i.e., assigned a number) by the order they are added and can be removed to delete the filter. Additional functionality is provided to combine the search results from multiple tables. This is provided through a logical dropdown menu, with the following options: 1) “AND”– the intersection of reports from the current and previously defined filter, 2) “OR”– the union of reports from the current and previous filter, and 3) “NOT”– the set difference of reports from the current and previous filters. A list of available filters is made available to the user with along an editable logic statement that specifies the combined search criteria, generated from the initial logical dropdown menus (e.g., ((1 AND 2) OR 3) represents the union between filter 3 and the intersection between filters 1 and 2). This logical statement can be adjusted based on any possible combination/permutation of filters– for instance, ((1 AND 3) OR (2 AND 4)), representing the union between the intersection between filters 1 and 3 and separately the intersection between filters 2 and 4. Once the final conditional logic has been specified, physicians can select one or more target table(s) for viewing. Running the final query will generate the final set of pathology reports selected using the conditional logic states and merge together the selected tables by these unique identifiers. Further description of the search process can be found in the supplementary materials (**Supplementary Figure 1**).

### Interactive Result Table

The final table display is visualized in the lower right of the web application, generated after running the query (**Figure 3**). The number of unique patients / reports can be found using the “Table Rows Summary” button. The final table is organized into multiple pages. Sorting and filtering operations, similar to that featured in an excel spreadsheet, can be used to further sort, query or subset the table by multiple columns. All the results can be exported/downloaded into multiple spreadsheet formats for further analysis (**Supplementary Figure 2**).

**Figure 3:**
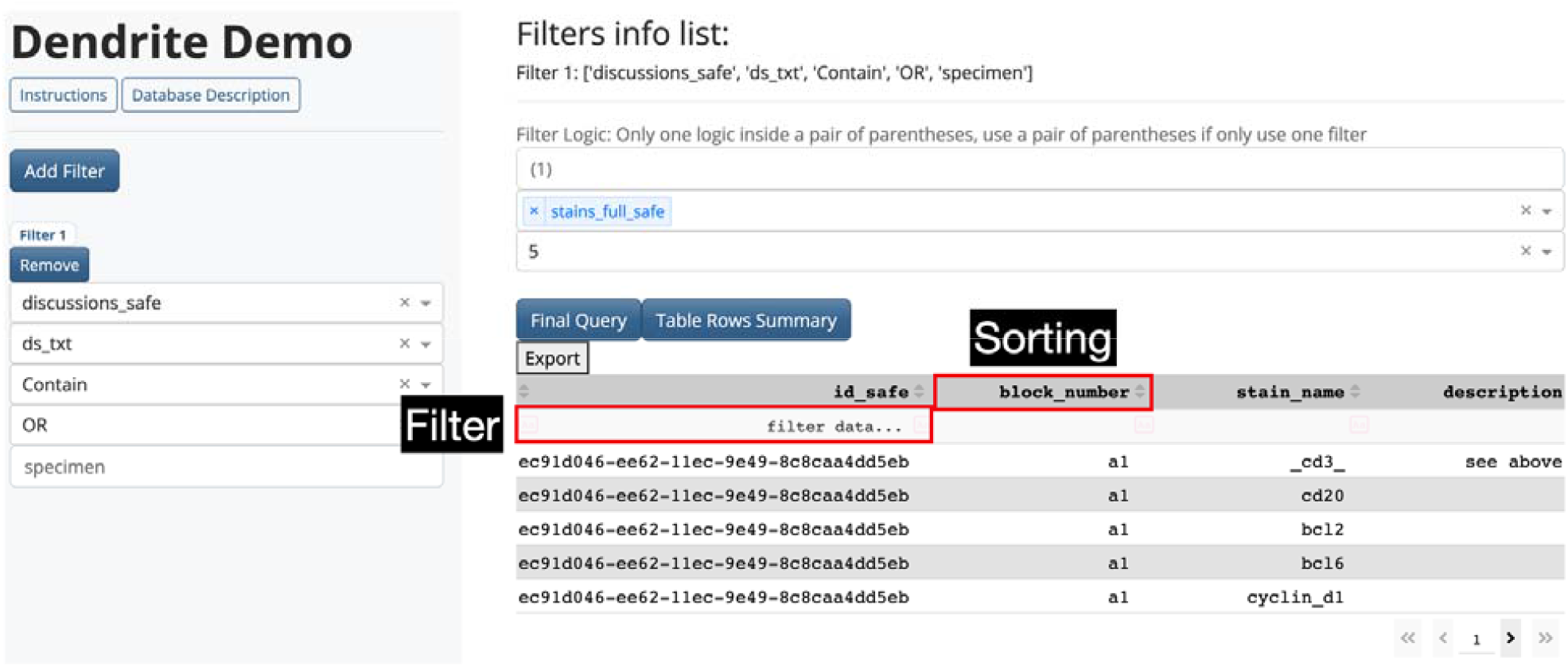
Illustration of filtering and sorting functionality in results display table Results.

### Description of Select Database Tables

A total of 749,136 reports were extracted from December 2011 to December 2021 (**Figure 4**), corresponding to 272,714 patients, assessed across 13 diagnostic subspecialties from 79 pathologists. Of these cases, 46,846 required intradepartmental consult (extraction of “qacc” from report), leading to a total of 2,764 consensus conferences (extraction of “conf”) and discussion amongst pathologists were reported for 3,330 reports (“dw_doc” reporting signature). Synoptic reporting was identified in 9,700 reports, though the availability of this information continues to grow. Addendums were found for 26,026 reports. Staining results were reported across 165,140 specimen blocks, and reports were linked to a total of 388,679 whole slide images. Clinical history, nuanced molecular findings (e.g., MLH1 hypermethylation), and flow cytometry findings were reported for 494,122, 162,059, and 3,065 reports respectively.

**Figure 4:**
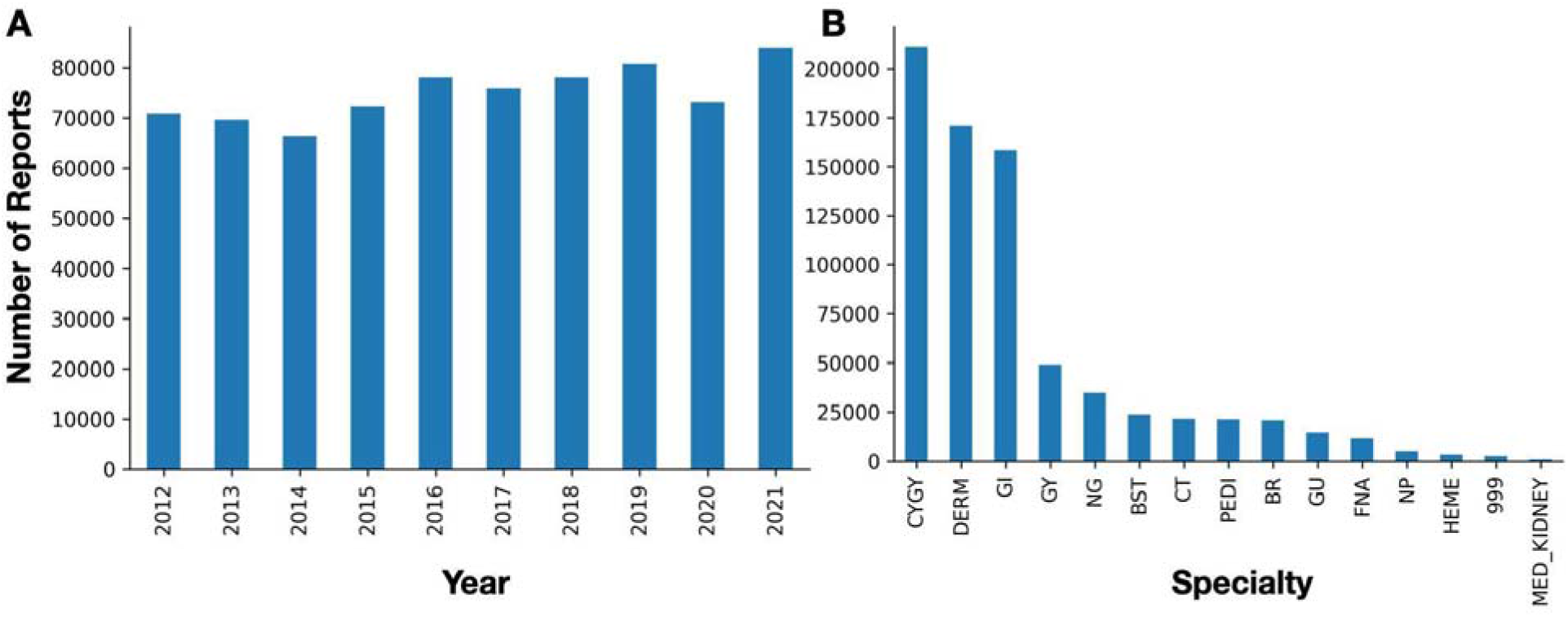
Breakdown of number of included pathology reports by: **A)** Year, **B)** Subspecialty

### User Tests

As an initial test of Dendrite’s functionality, we asked collaborating pathologists to perform a series of example queries. Future works will explore the impact this database system has on facilitating timely review by assessing widespread usage across the department but was outside of the scope of this technical note.

#### Urine Specimen Atypia

A cytopathologist conducted a search to locate voided urine cytology specimens to assess instances of specimen atypia for the assessment of high-grade urothelial carcinoma. First, cases were filtered using the non-gynecological specimens table (*ng_safe*), searching for voided urine in the specimen *source* field (**Figure 5**). While we were able to identify a number of specimens using this search functionality, ultimately we opted to use the diagnostic text (*diagnoses_safe*), searching for voided specimens, as we felt the free text was less restrictive (**Figure 6**). By specifying a search for voided specimens, we were able to remove washes and upper tract specimens. This search yielded approximately 4,178 reports, close to the number of voided urine specimens reported over the data collection period. We queried the diagnostic tables and merged these with a table pertaining to additional pathology reporting information, including sign-out time. This table was exported to a CSV format and further processed using a custom R script (using the “grepl” function) to yield the number of negative, atypical, suspicious and positive cases over time. We plotted the incidence of these diagnostic categories over time along with a barplot depicting the overall categorization in **Figure 7**, demonstrating that in less than a minute, we could conduct a longitudinal study of urine specimen atypia to inform rapid bladder cancer screening. For instance, by plotting the incidence of different diagnoses, we found that the number of negative findings increased from 2016 to 2018, while the number of positive findings decreased from 2016 to 2018. As our department implemented The Paris System for Reporting Urine Cytology in 2018 ^14^, these findings corroborate with previous research indicating early adoption of these reporting guidelines. Further analysis was conducted using hierarchical Bayesian regression modeling (utilizing the *brms* and *emmeans* R packages) ^15–17^. In this model, we treated the level of implementation—pre-publication (before 2016), publication of the guidelines prior to its actual implementation (2016-2016), and post-implementation (after 2018)—as an ordinal variable with monotonic effects. Incorporating pathologist-level random intercepts, we identified a reduction in specimens deemed atypical across the implementation stages, as detailed in **Table 1**.

**Figure 5:**
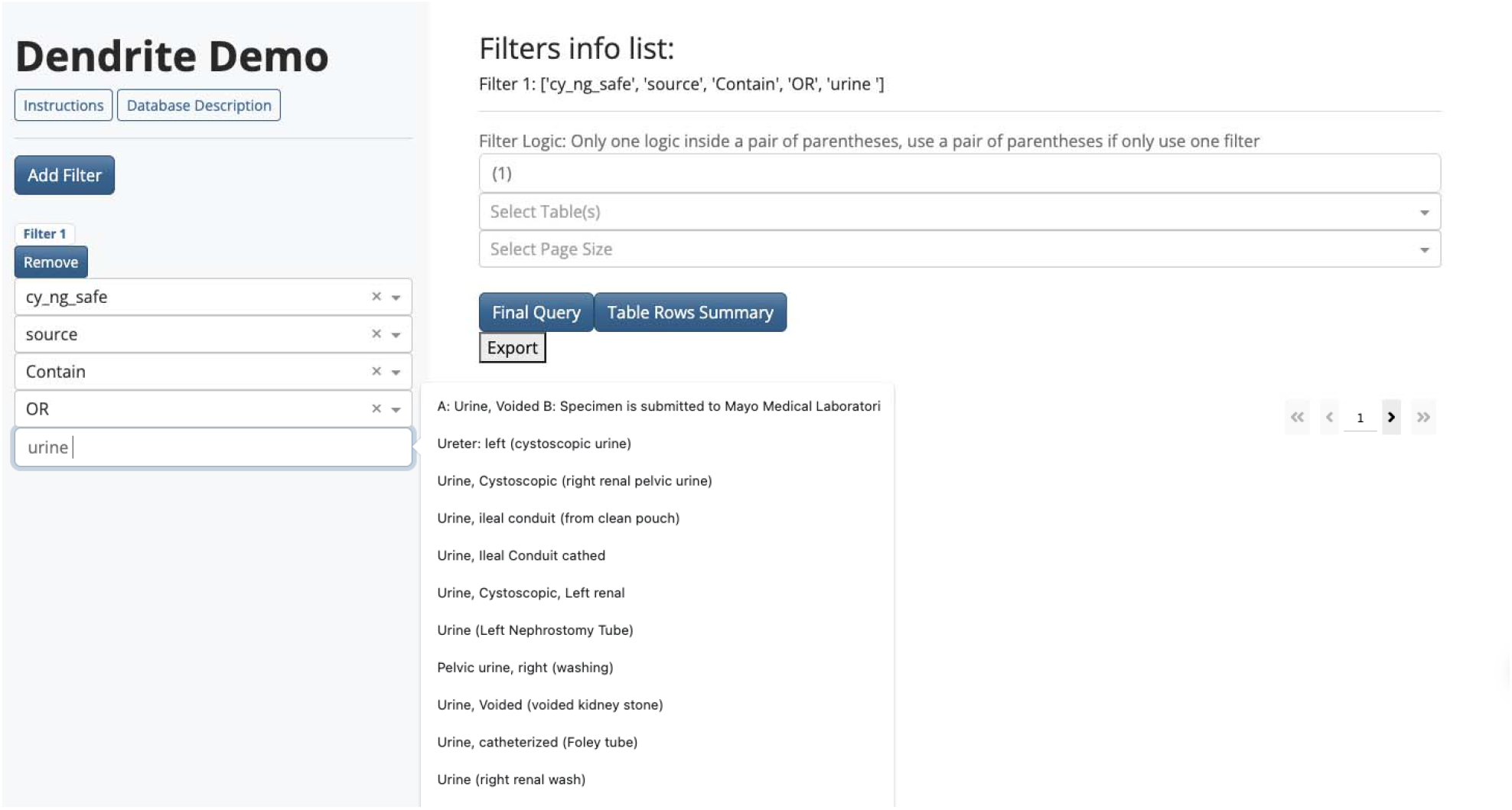
Querying non-gynecological table for voided urine specimens.

**Figure 6:**
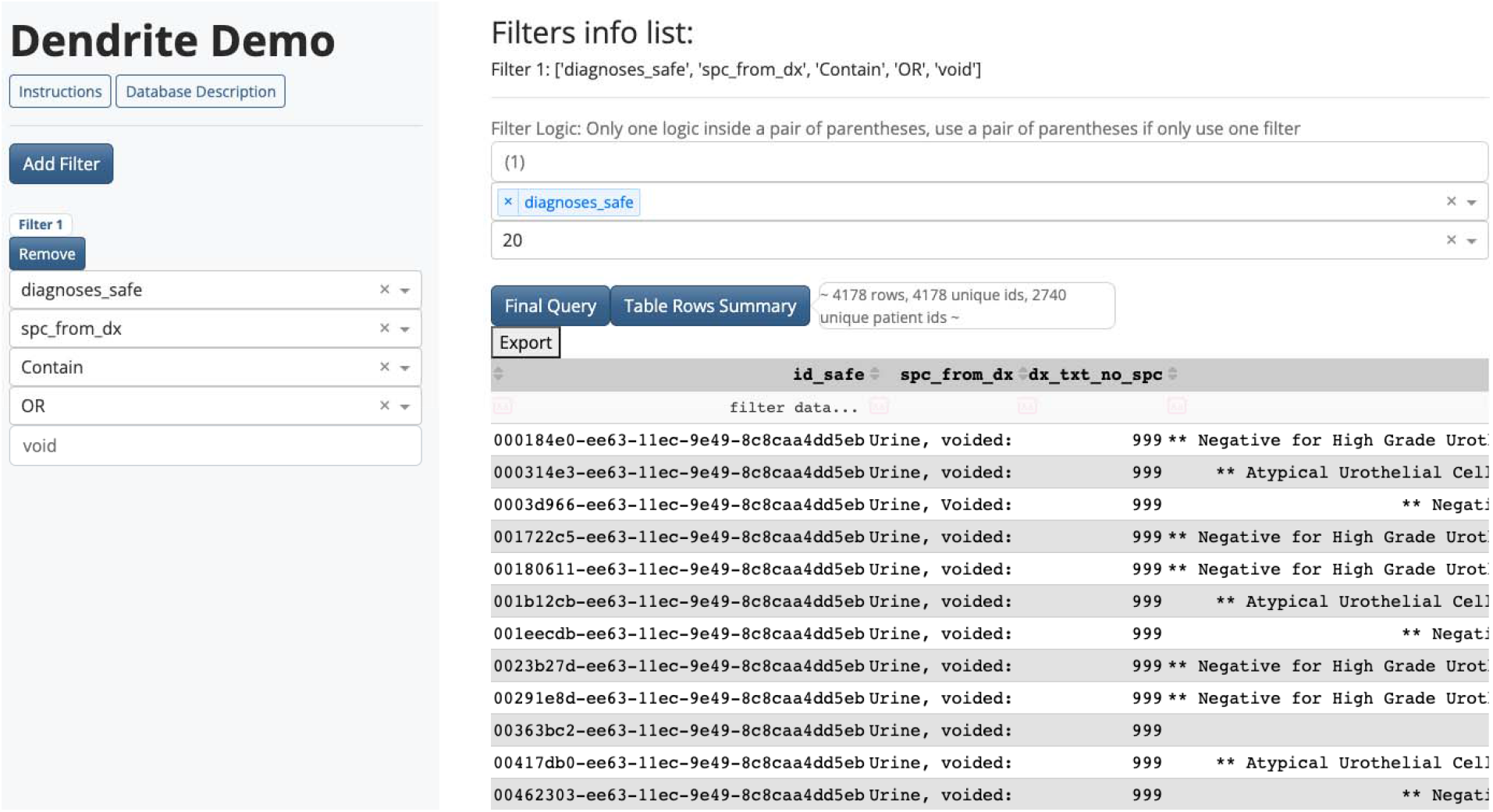
Querying diagnostic text for voided urine specimens.

**Figure 7:**
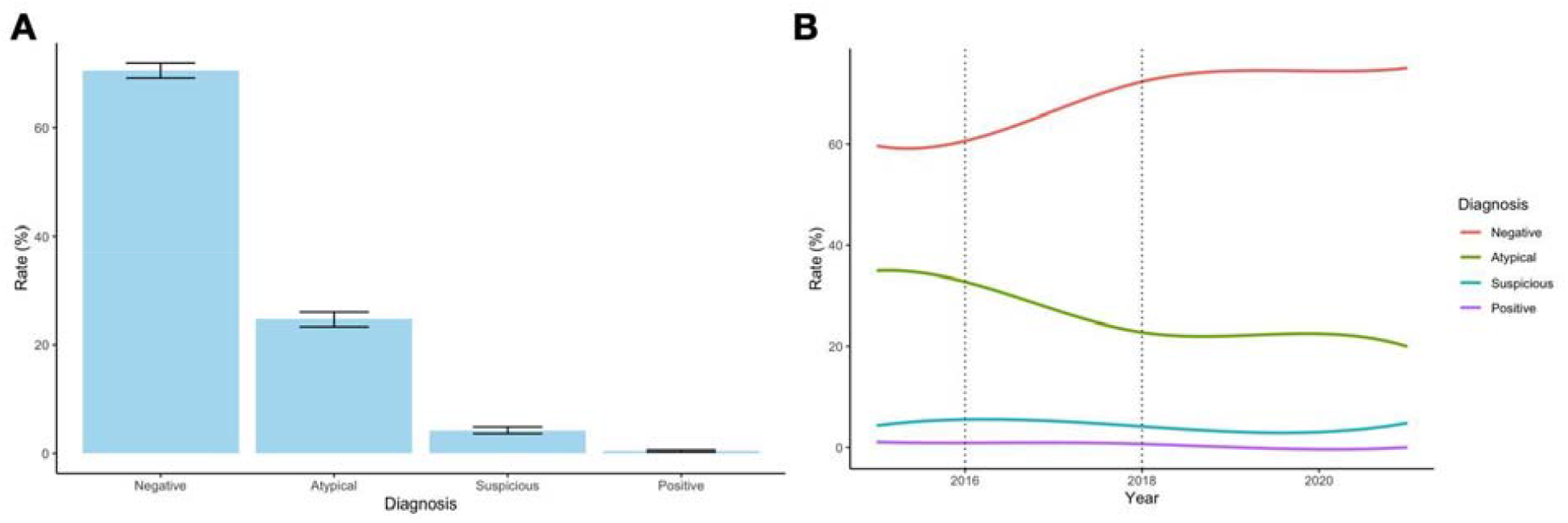
Breakdown of The Paris System diagnostic assignments: **A)** Aggregated across the entire database reporting period, and **B)** Yearly over time across the reporting period; vertical lines indicate publication of the official TPS guidelines (2016) followed by official implementation of the guidelines in 2018

**Table 1:** Regression Coefficients from Statistical Model Comparing Atypia Rates between Various Implementation Periods for The Paris System: Odds ratio above one indicates that atypia rates increased over time whereas below one indicates reductions in reported atypia compared to the other TPS categories

| Parameter | Odds Ratio | 2.5% CI | 97.5% CI | p-value |
| --- | --- | --- | --- | --- |
| Degree of Implementation: Post-Implementation > Post-Publication > Pre-Publication | 0.90 | 0.82 | 0.97 | 0.01 |
| Post-Implementation vs. Post-Publication | 0.94 | 0.80 | 1.00 | 0.01 |
| Post-Implementation vs. Pre-Publication | 0.80 | 0.68 | 0.95 | 0.01 |
| Post-Publication vs. Pre-Publication | 0.87 | 0.72 | 0.99 | 0.01 |

#### Colon adenocarcinoma cases

A GI pathologist attempted our second search. We sought to pull MLH1 staining results from colon adenocarcinoma cases to assess for mismatch repair deficiency to build a retrospective cohort that assesses the metastatic potential of this subgroup in addition to pulling corresponding whole slide images for a digital image analysis. To do this, three filters were constructed– the first two from the diagnostic text tables and the final one from the staining results tables. We first filtered by whether the text contained “colon” and “adenocarcinoma”. The final filter sought to query for cases where MLH1 staining had been done. Multiple stains were ordered for specific cases. The interactive filtering mechanisms of the final display table were used to subset stains by MLH1 status. For the final query, this table was merged with a whole slide image table, which specifies the file locations of all matching whole slide images for tissue slides stained with hematoxylin and eosin within our Aperio Image server. This search yielded approximately 676 pathology reports (**Figure 8**). We conducted additional filtering for other mismatch repair related genes (MLH1/PMS2 and MSH2/MSH6 form two separate heterodimers with different progression characteristics but indicative of the microsatellite instability pathway). Results were exported to a CSV file. The CSV file contained information on staining results and the file locations of whole slide images within our Aperio image server. Staining results were further processed using a custom R script to generate a bar plot of negative and positive staining findings, identifying significantly higher instances of loss of expression for the MLH1/PMS2 heterodimer as assessed through immunohistochemistry (**Figure 9A**). This information was also used to pull the corresponding whole slide images of H&E stained tissue (**Figure 9B**).

**Figure 8:**
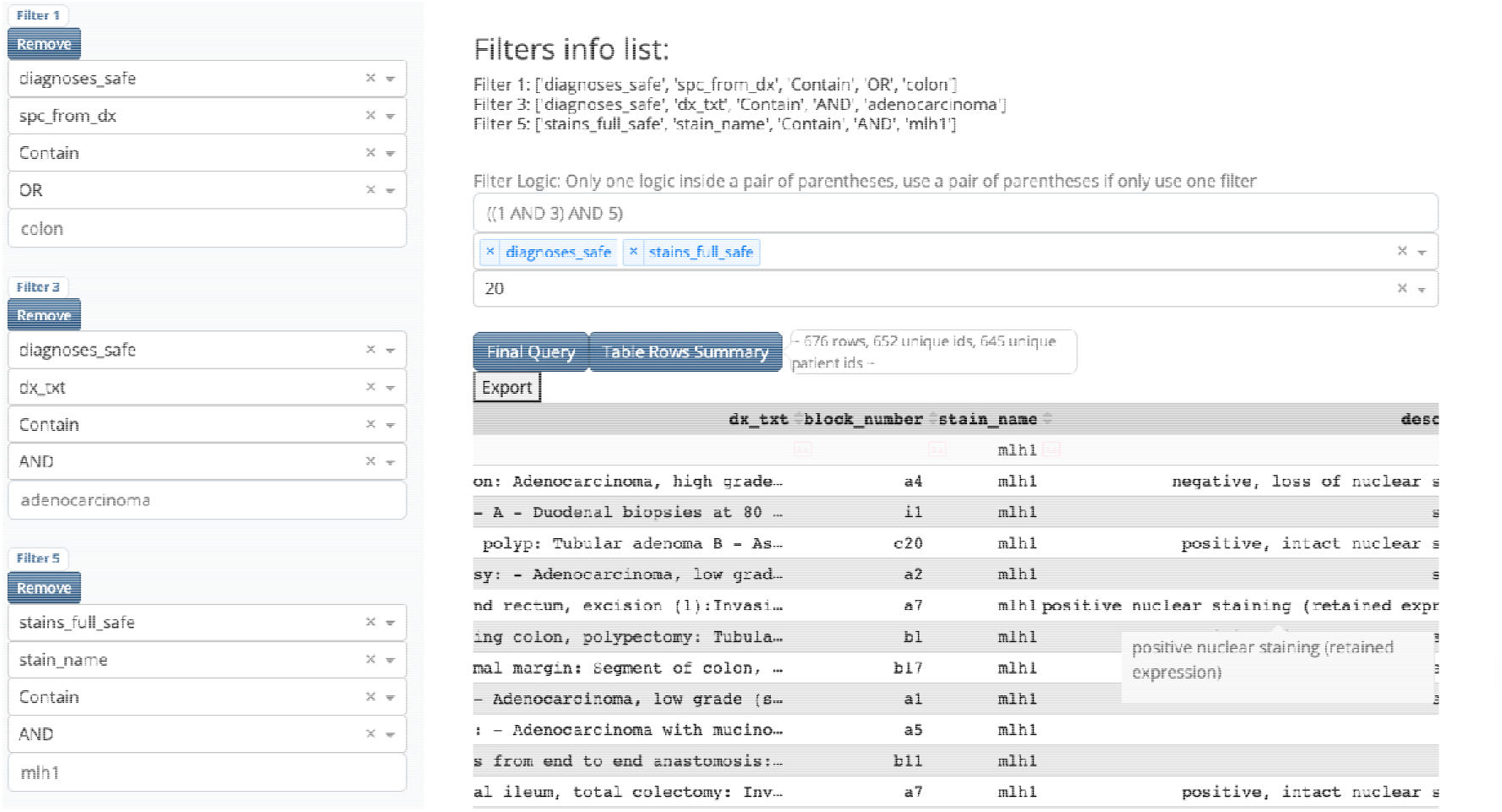
Query results of diagnostic text and staining information for Colon Adenocarcinoma cases for potential mismatch repair deficiency.

**Figure 9:**
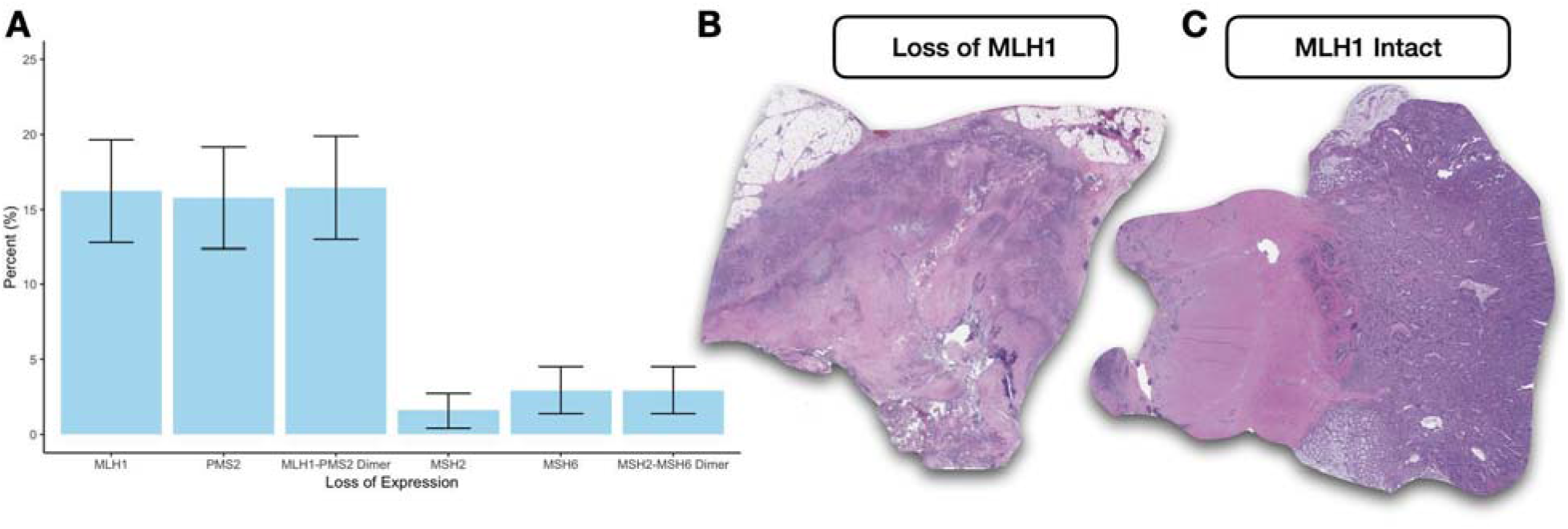
Further analysis of extracted Dendrite table for studying colon cancer mismatch repair deficiency: **A)** Breakdown of loss of expression for mismatch repair genes as assessed through immunohistochemistry; **B)** Whole slide images extracted using Dendrite image file location information illustrating slides with and without MLH1 positive staining

#### Qualitative description of the pathologist experience using Dendrite

From discussions with pathologists in our department, we found that compared to the search functionality from our EMR (Cerner), the pathologists found Dendrite to be faster, more responsive and flexible. Pathologists were able to finalize search criteria across hundreds of thousands of cases and merge information from disparate data sources in seconds. The secondary search / filtering capabilities offered using the interactive display table were found to be particularly effective in further refining the search results.

## Discussion

By leveraging expert domain knowledge and natural language processing algorithms to parse free-text pathology reports into structured multi-table formats, Dendrite presents a viable database tool that pathologists can use to rapidly build study cohorts and leverage for quality improvement purposes to complement other emerging EMR systems.

While we have implemented an initial prototype of the Dendrite web application and deployed this tool over AWS, there are many areas where this tool can be improved. First, we plan to develop and expand a user management system, which will allow for user logins and permit tracking of user data ^18^. This will help us determine the broad utilization of this tool within the department and areas to improve this application for further widespread adoption. Users will be able to access their search history offline to help guide more informed searches as users gain more familiarity with the system. Leveraging the users’ search history can also help personalize further searches based on their preferences (e.g., accurate suggestions and operations). For example, if some users are accustomed to querying data about colon cancer, then the keywords related to colon cancer can be suggested through the integration of knowledge and semantic databases which link biological entities. Natural language processing algorithms can continue to improve their suggestions and complex instructions through the accumulation of more data and user feedback.

Furthermore, we plan to further integrate NLP (natural language processing) and machine learning technologies into this web application in order to mine free text data to extract structured reporting information that may be more readily queryable. This will be accomplished by configuring multiple machine learning algorithms to make predictions across multiple tasks, including but not limited to 1) CPT code prediction to identify instances of underbilling ^19^, 2) named entity recognition tasks (e.g., extraction of staining information, histological findings) ^20^, and 3) instances where an outside consult is needed, etc. Trained predictive algorithms can be further integrated into future iterations of this web application. large language models (LLMs) offer a promising approach by capturing nuanced contexts and relationships. We acknowledge that the reporting information is still incomplete and biased towards certain subspecialties, and we are aware that more work is needed to extract information from less common or structured specialties. This database features record linkages to imaging data. Further integration of our text database into our genomics and imaging infrastructure can enhance its search capabilities as well as motivate future multimodal analysis ^21^.

## Conclusion

Dendrite, a web application developed using natural language processing and expert knowledge, offers pathologists a flexible search capability for conducting large-scale assessments of clinical pathology reports in the context of clinical research and quality improvement. The application employs various features to enable efficient search functionality, including filters that can be combined using conditional logic statements, comprehensive merging of reporting tables associated with selected pathology reports, and dynamic sorting and filtering operations on the display table. These features greatly facilitate the search process for pathology data, making it easier and more convenient for pathologists in their clinical and research endeavors.

## Data Availability

All data produced in the present study are available upon reasonable request to the authors

## Supplementary

**Supplementary Figure 1:**
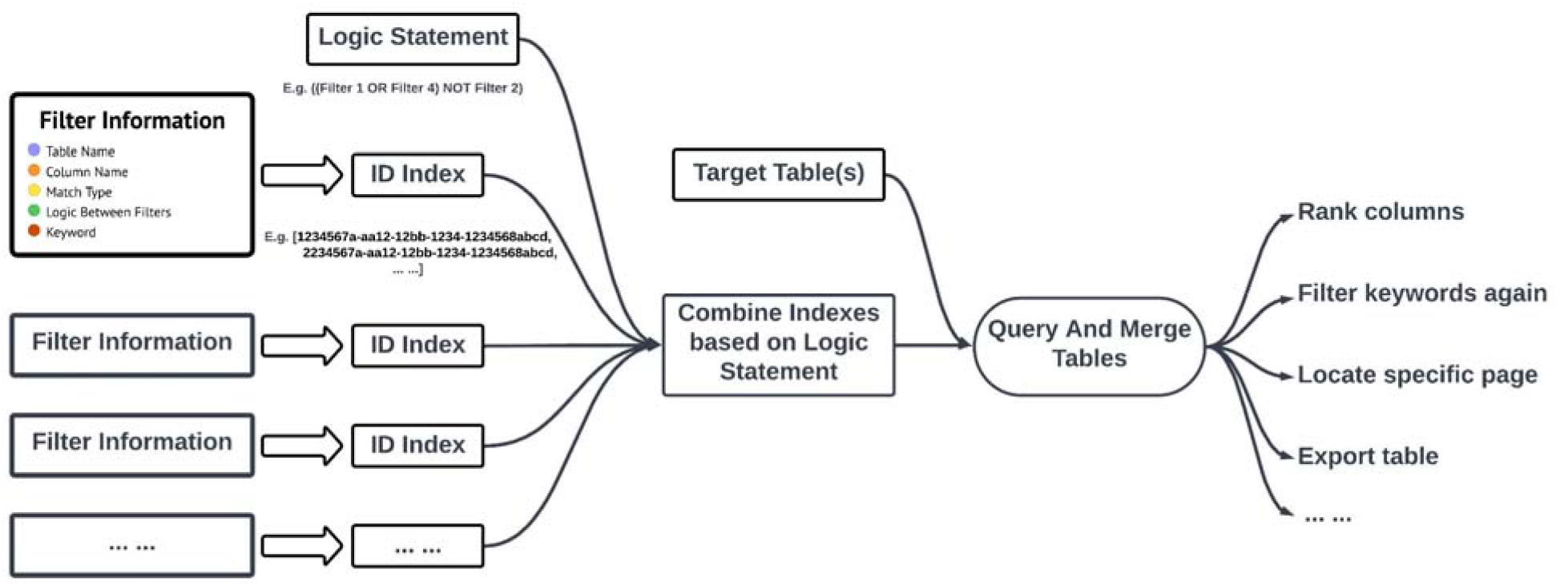
Illustration of table querying logic using Dendrite Web Application: Separate filters are specified by the user. Each filter returns a list of pathology report IDs. Indices are joined through intersection and union operations based on user-specified logic statements. Target tables are selected, each filtered based on the final set of report IDs. Additional table operations may be performed prior to export.

### Additional Information on Dendrite Web Application Text Search Workflow

In the intermediate level of the Dendrite searching process, every action on the interface level will have a corresponding processing interaction in the underlying level. The first step is to collect the information from the filters, including table name, column name, match type, logic choice and keyword. It involves multiple callbacks. For example, the column name depends on the table. The column choices will only update after the table is chosen. Each filter will generate an index list from a specific table. All of the filters’ information will be displayed on the right side of the screen, so that it remains visible to facilitate user experience. Filters are removable, and once removed, will be deleted from the display window with no information saved.

The next step is to verify the logic statement. By inputting logic choices in the filter, Dendrite will automatically generate a logic statement in the order of the filter indexes. Physicians can also edit the logic statement by adding, removing and reordering the filter indexes. Dendrite will then parse the statement and query the index in order. It will then combine the indexes based on the logic between them. For example, if the logic between two filters is “OR”, Dendrite will keep all indexes. However if the logic is “AND”, Dendrite will only keep the intersection of the indexes, and then form a final index list. After that Dendrite will extract the table, based on the target table(s) input by the physicians. It will retain the rows which have the same indexes as the final indexes list. And then merge these tables together to the final table. During the merging, if a missing value appears, it will be left blank. Clicking on Query will display the Table. Inside this table, physicians can further filter the keyword or perform operations. Dendrite will recognize the column content type as a string data or numeric data, and then rank it in alphabetical order or numerical order. And the table data will be saved. The generated table data will be exportable via the download option.

Physicians can select multiple keyword-based search terms at the same time and combine these search terms in any way they want. For example, they can select any two conditions that are logically related to “AND” or “OR” or “NOT”. This allow physicians to perform real-time interactive data manipulation across one or more target tables that have been merged together. Also, the application automatically recognizes the table data types, allowing it to perform different sorting operations for tables that contain character and numeric data. Physicians can filter the table further by adding filters. While taking into account the readability of the data, the data table provides page numbering and scrollable axis functionality. This allows for more precise targeting of rows and also extends the table’s display range while maintaining readability.

**Supplementary Figure 2:**
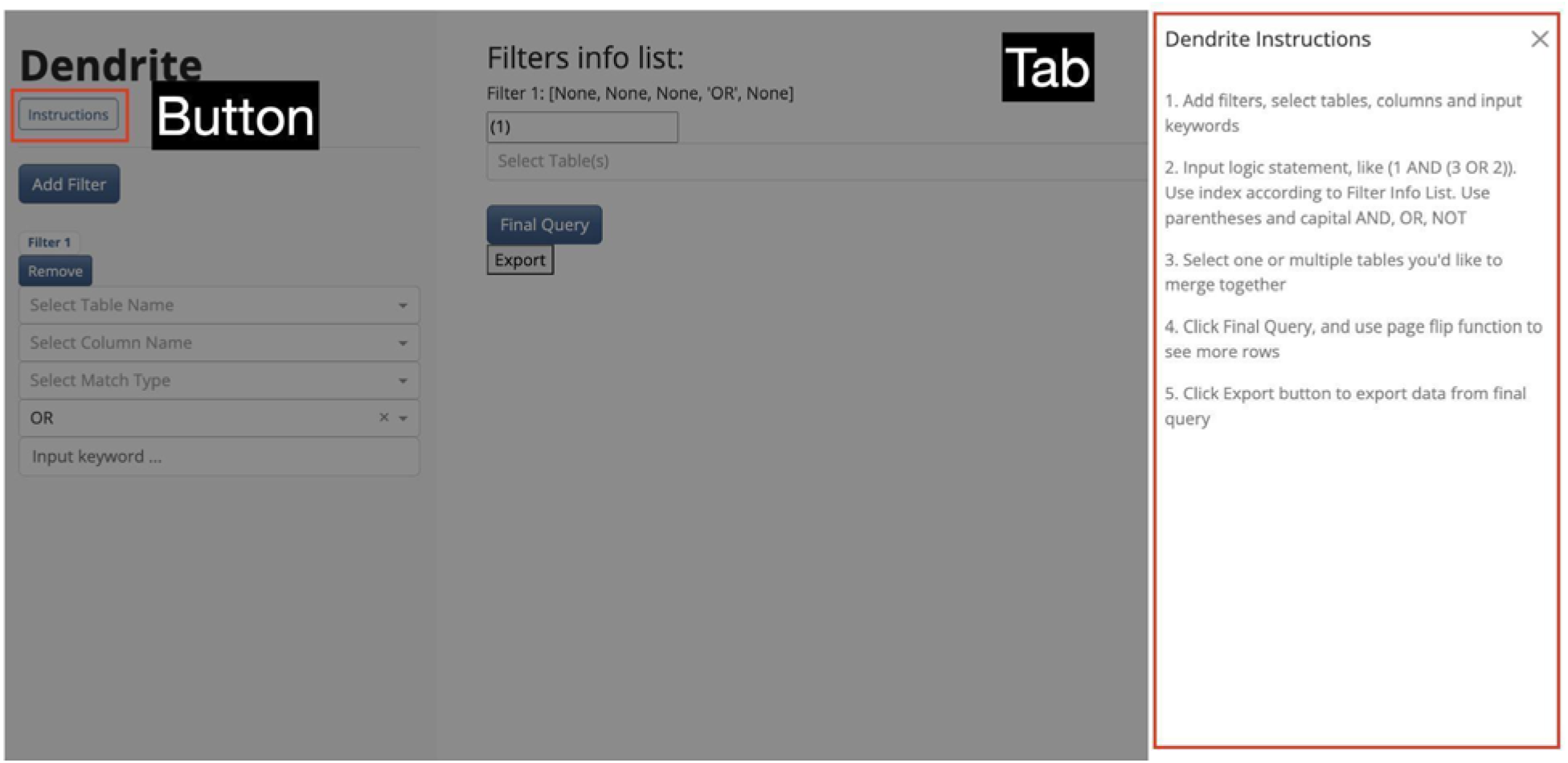
Dendrite Instructions Tab. In case of unfamiliarity with operation of the web application, we developed an instruction tab that will appear by clicking the instruction button directly below the title. With readability in mind, we designed the instruction tab to appear by sliding from the right. This allows users to read the guide while simultaneously having access to the main dendrite content on the left, enabling immediate recognition on the interface of salient features described in the instructions.

### Additional Information on Structured Dendrite Table Schema

**Supplementary Table 1:**
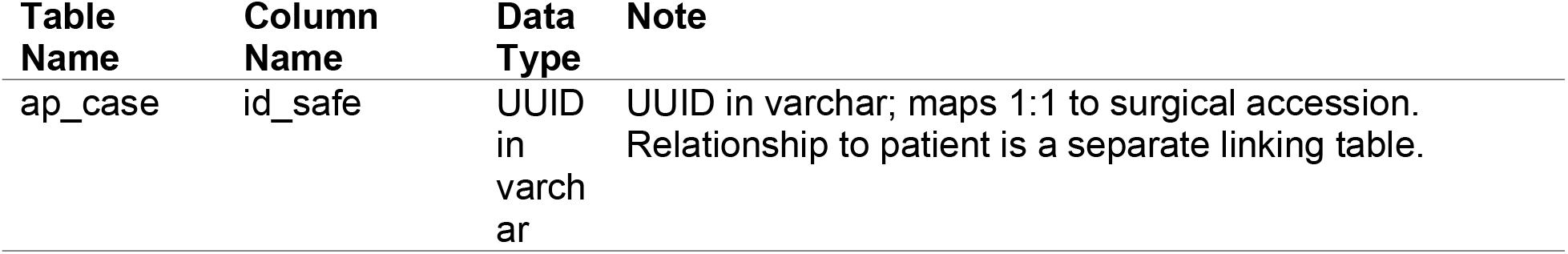
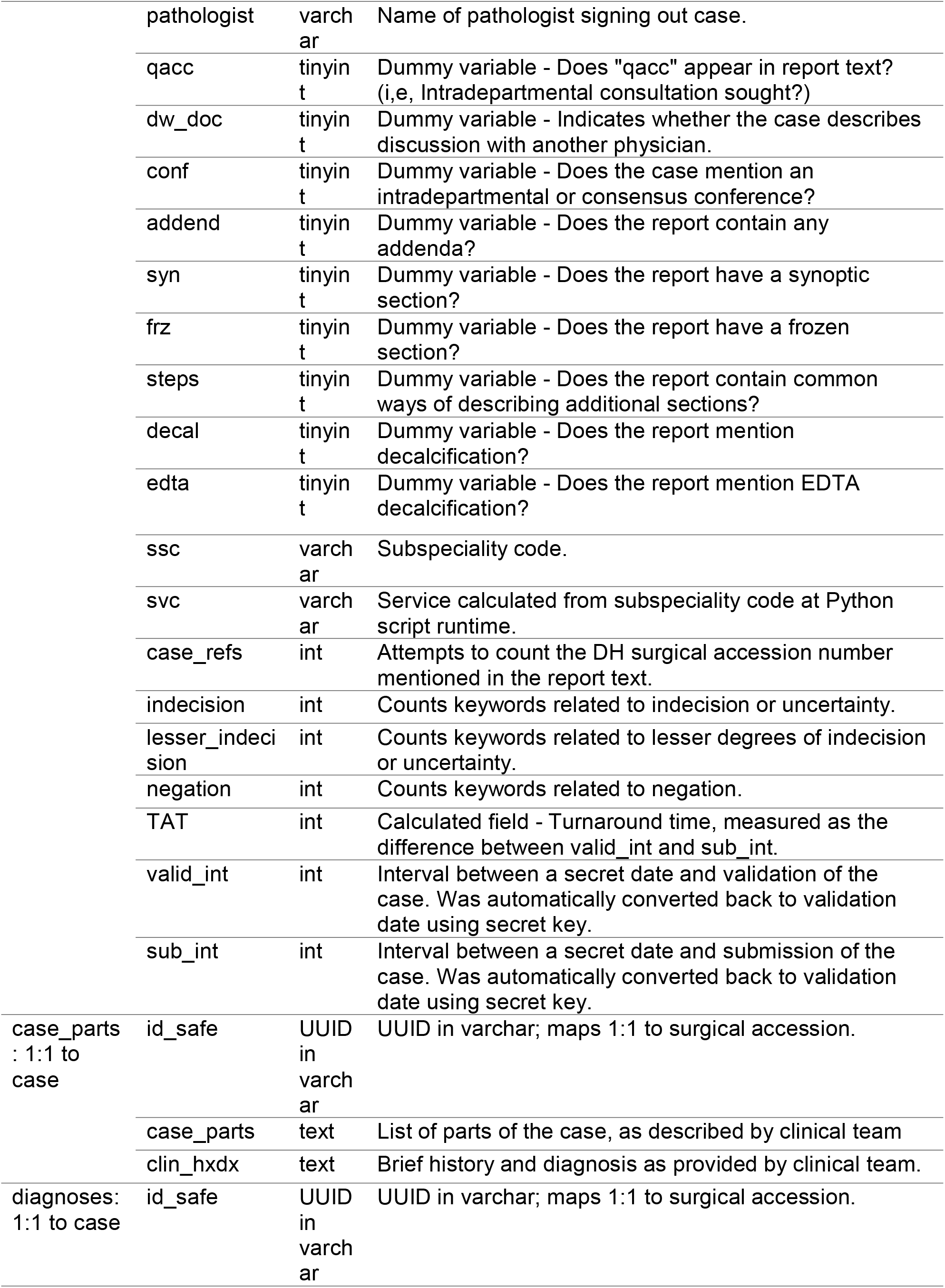
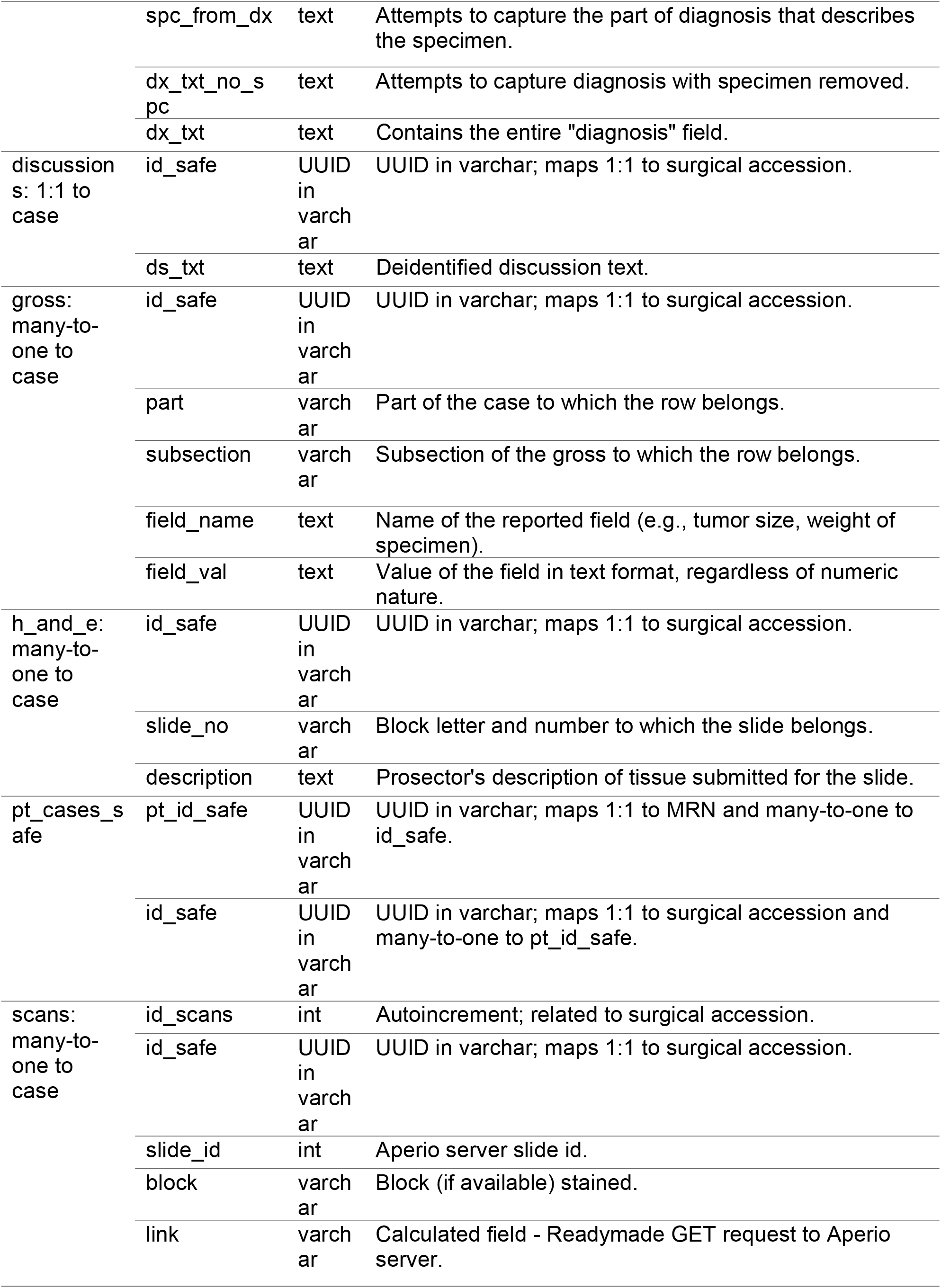
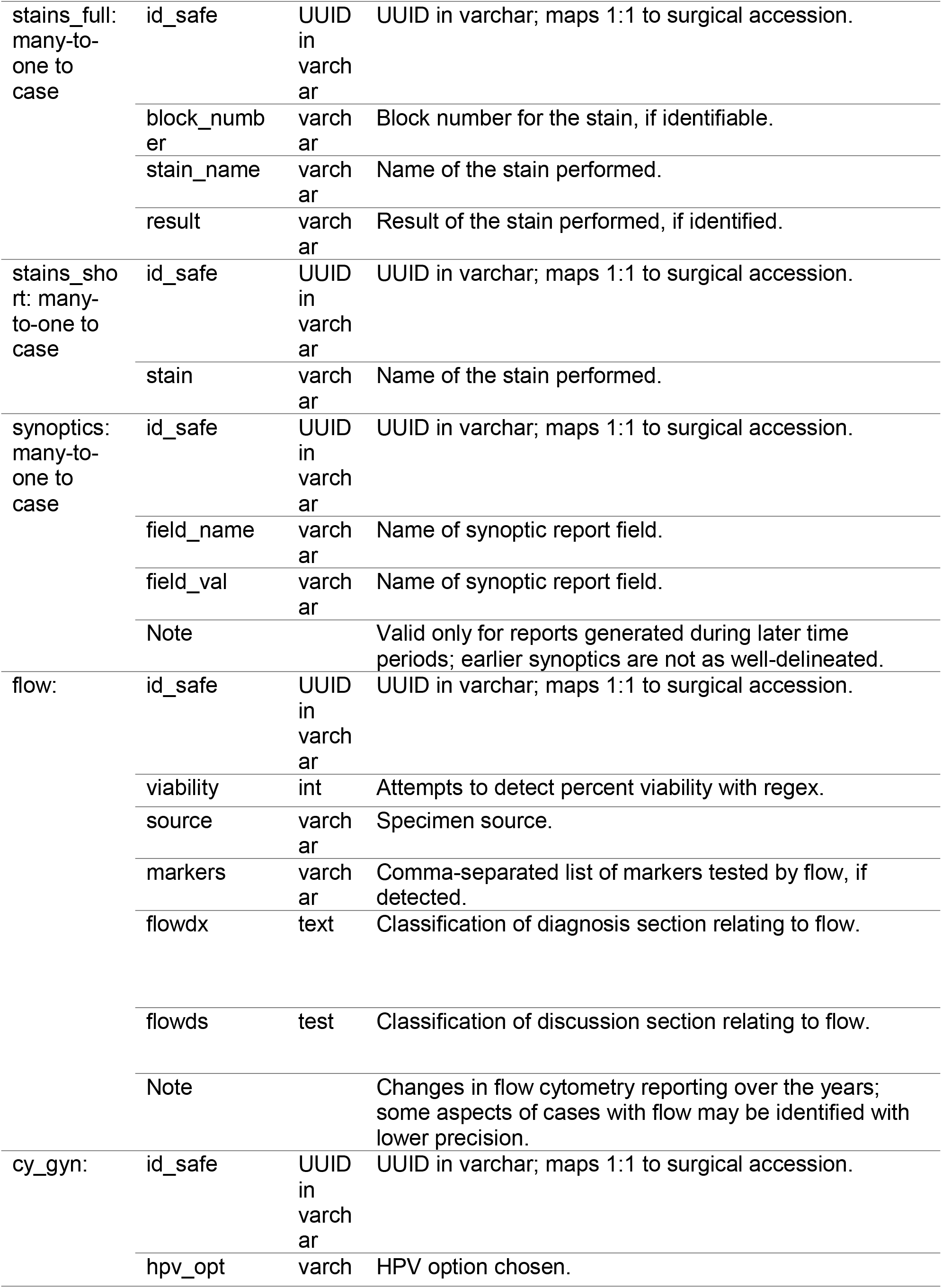
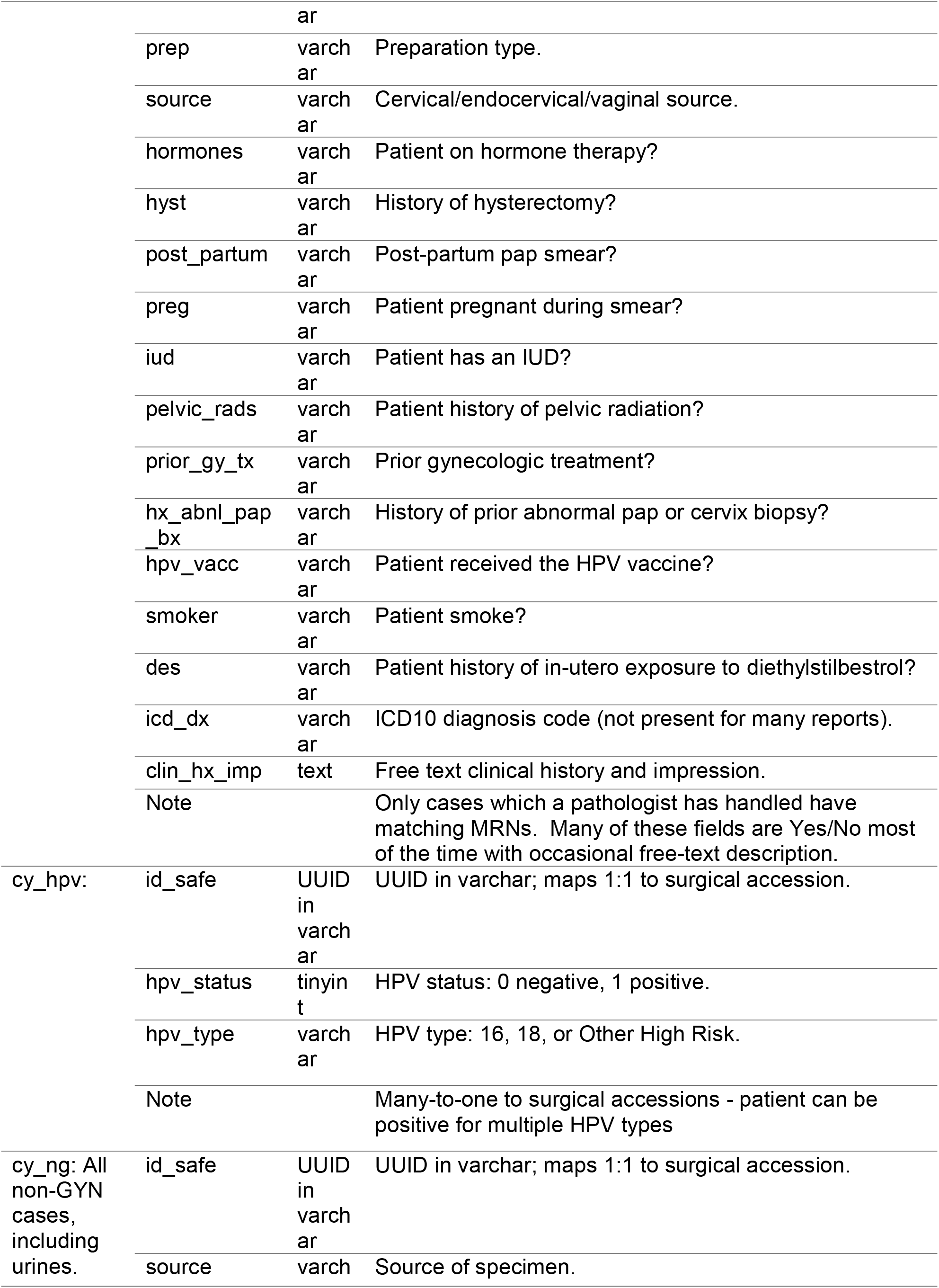
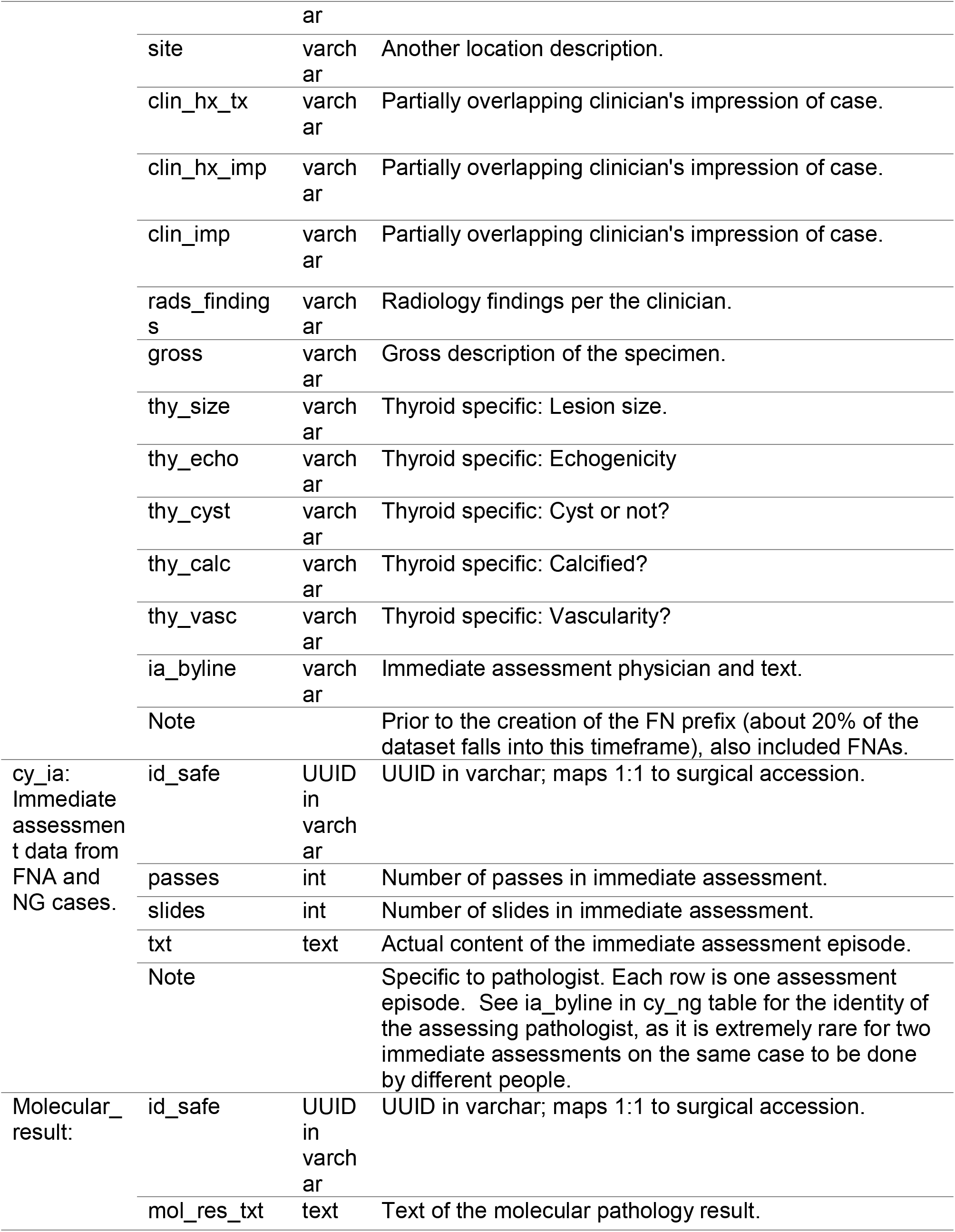
Pathology database table names, column names, data types and metadata

